# Assessing the utility of Fronto-Parietal and Cingulo-Opercular networks in predicting the trial success of brain-machine interfaces for upper extremity stroke rehabilitation

**DOI:** 10.1101/2025.04.08.25325026

**Authors:** Gokul Krishna Raja Padmaja, Nikunj Arunkumar Bhagat, Pragathi Priyadharsini Balasubramani

## Abstract

For stroke participants undergoing motor rehabilitation, brain-machine/computer interfaces (BMI/BCI) can potentially improve the efficacy of robotic or exoskeleton-based therapies by ensuring patient engagement and active participation, through monitoring of motor intent. In such interventions, exploring the network-level understanding of the source space, in terms of various cognitive dimensions such as executive control versus reward processing is fruitful in both improving the existing therapy protocols as well as understanding the subject-level differences. This contrasts to traditional approaches that predominantly investigate rehabilitation from resting state data. Moreover, conventional BMIs used for stroke rehabilitation barely accommodate people suffering from moderate to severe cognitive impairments.

In this first-of-the-kind study, we explore the cognitive dimensions of a BMI trial by probing the networks that are core to the BMI performance and propose a network connectivity-based measurement with the potential to characterize the cognitive impairments in patients for closed-loop intervention. Specifically, we tease apart the extent of cognitive evaluation versus executive control aspects of impairments in these patients, by measuring the activation power of a major cognitive evaluation network-the Cingulo-Opercular Network (CON) and a major executive control circuit-the Fronto-Parietal network (FPN), and the connectivity between FPN-CON. We test our hypothesis in a previously collected dataset of electroencephalography (EEG) and structural imaging performed on stroke patients with upper limb impairments, while they underwent an exoskeleton-based BMI intervention for about 12 sessions over 4 weeks. Our logistic regression modeling results suggest that the connectivity between FPN and CON networks and their source powers predict trial failure accurately to about 84.2%. In the future, we aim to integrate these observations into a closed-loop design to adaptively control the cognitive difficulty and passively increase the subject’s motivation and attention factor for effective BMI learning.

## 1. Introduction

Loss of motor function is prevalent in most post-stroke patients and significantly impacts their activities of daily life (ADL) (Gilmore & Spaulding, 2001). A brain-machine interface (BMI) has been shown to be a promising rehabilitation system for people with severe motor disabilities (Ang et al., 2015; Biasiucci et al., 2018; Buch et al., 2008). It operates by sensing desired brain activity patterns to control an external device that can assist with restoring movement by enhancing the sensory, motor, and cognitive associations between the desired and executed action, thereby providing neurofeedback and promoting neuroplasticity (Daly & Wolpaw, 2008; Soekadar et al., 2015).

Mental imagery and practice improve the condition of affected limbs in stroke patients (Page et al., 2005). The ability to imagine making a movement is intact in such patients, therefore using those intention signals to make an external device move to support their action and improve their condition was promising and incorporated in many studies (Buch et al., 2008; Ramos-Murguialday et al., 2013). For example, (Frolov et al., 2017) showed significant improvement in upper-limb functionality in stroke patients who went through a repeated imagery-based BMI protocol. They trained a classifier using imagery-related EEG signals and used that classifier-exoskeleton set up to support some simple hand movements for the patients. A study by (Bhagat et al., 2020) presented such a classifier that acquired up to 79±18 % accuracy in classifying motor intention showing the potential advantage of BMI-based rehabilitation protocols.

While BMI-based neurorehabilitation has been shown to be effective in several clinical trials, earlier studies rely more on resting state data and haven’t well explored the network-level mechanisms, in terms of various cognitive dimensions such as executive control versus reward processing for controlling the system processes. Moreover, conventional BMI training isn’t adaptive enough to compensate for patient fatigue, memory load, and other cognitive distractors, which often make the sessions demotivating and even insurmountable for many patients. In an age of more computational power along with the established neuroscientific theories on human cognition, it is time to make such interfaces able to pay heed to the cognitive states of the patient. Though earlier studies explored the pre-BMI neural patterns extensively in terms of decoding the preparatory motor activations and precise motor actions for controlling the external device, the use of reward information within BMI paradigms is still early in development (Khajuria et al., 2024; Mitra et al., 2023; Soriano-Segura, Ferrero, Gracia, et al., 2023; Soriano-Segura, Ferrero, Ortiz, et al., 2023).

Understanding reward processing can inform about the motivation of the participant in performing the BMI task, and importantly the error processing ability of relevance to their learning, both of which are suggested to significantly contribute to the BMI performance. There could be at least two networks in the brain of significant importance to motor execution, learning, and eventual motivation—the Fronto-Parietal (FPN) and Cingulo-Opercular networks (CON). The FPN activations are suggested to be dominantly encoding motor preparation, intent, and motor action in BMI (M. Liu & Ushiba, 2022), while the CON activity is suggested to dominantly encode error-related processing, outcome learning, and cognitive control (Soriano-Segura, Ferrero, Gracia, et al., 2023; Soriano-Segura, Ferrero, Ortiz, et al., 2023).

In this study, we specifically asked whether brain network activations can predict the success of the ongoing trial and whether it is the pre-BMI response or the post-BMI response neural activation, suitable for understanding reward processing following a successful BMI decision. We hypothesized that for a successful BMI trial, the cognitive and executive brain networks should communicate effectively, such that successful and unsuccessful movement predictions are appropriately reinforced. Further, we also hypothesized that the connectivity between FPN and CON will be weaker for unsuccessful BMI trials as compared to trials with successful BMI predictions. To prove our hypotheses, we analyzed data from ten stroke participants from a published study (Bhagat, et al, 2020) involving a BMI-based intervention for upper limb recovery, with simultaneously recorded electroencephalography (EEG) and electromyography (EMG) signals. In that study, patients had to make both a movement ‘intention’ that was detected by an EEG classifier and an ‘attempt’ that was captured by EMG signals to successfully drive the exoskeleton and accelerate the patient’s recovery. Here, we further probe the dataset, especially of the network activations in FPN and CON, to understand the BMI performance. We evaluate the network’s ability to offer a sneak peek into the outcome, in other words, the subjective value function associated with any trial, and their ability to *predict* the success of a BMI trial.

## 2. Materials and Methods

The aforementioned study (Bhagat et al., 2020) was conducted as part of a clinical trial (ClinicalTrials.gov #NCT01948739) and the data were collected from 10 chronic stroke patients. The patient demographics are provided in Supplementary Material A.

### 2.1. Experimental Set-up and Task

In (Bhagat et al., 2020), the MAHI Exo-II exoskeleton supported each patient’s impaired upper limb while they performed a simple reaching task, which focused on training their elbow flexion and extension. The trial began with a cue followed by a target that appeared at the top or bottom of the screen. Patients were told to think about performing the movement (i.e. “motor intention” phase) and then attempt the movement (i.e. “motor attempt” phase). The BMI algorithm used both EEG & EMG signals to validate the occurrence of signatures of intention and attempt and then initiate the exoskeleton’s movement (**Fig. 1A)**. Specifically, the algorithm used a nonlinear Support Vector Machine classifier that monitors the presence of intention through movement-related cortical potential in the delta band (0.1 – 1 Hz). After detecting intent from EEG activity, the BMI system monitored for subsequent EMG activity from the biceps and triceps of the affected arm to validate the attempt (using a standard deviation-based threshold value). If EMG activity corresponding to a motor attempt was not detected within 1 second of motor intent (predicted from EEG signals only), the exoskeleton wouldn’t move, and the trial would be labelled as unsuccessful. Therefore, patients had to perform both intention and attempt quickly and optimally to ‘successfully’ complete each trial. The trials were interleaved with randomized interval durations. The whole study spanned around 12 sessions for each patient (3x times a week for 4 weeks).

**Fig. 1.**
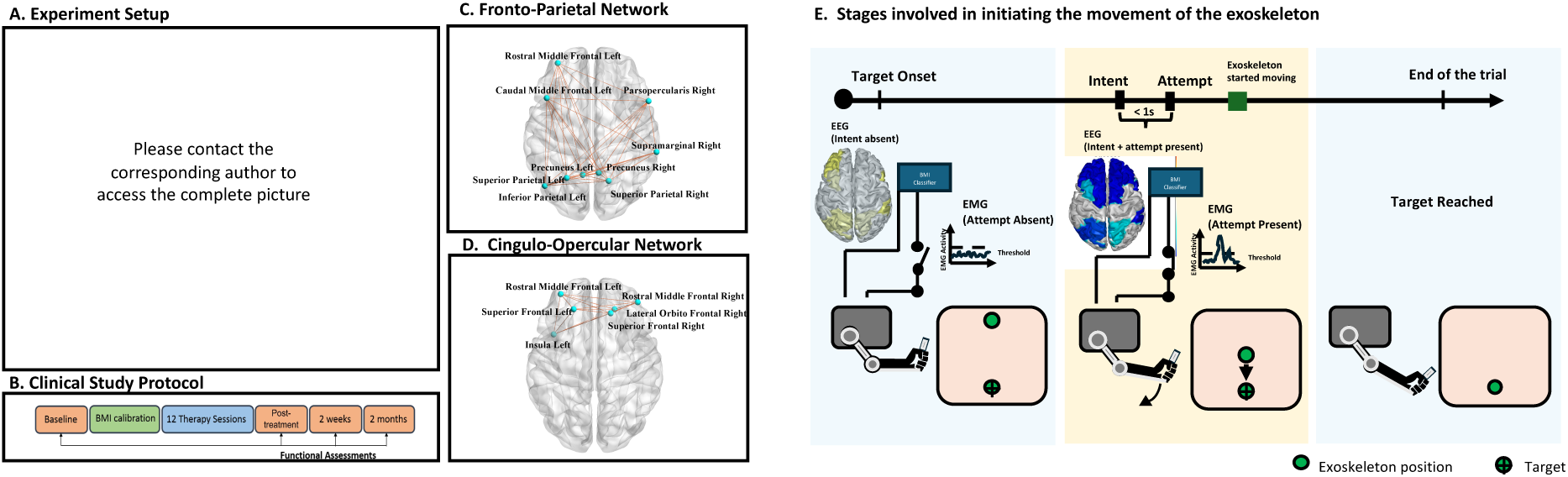
The BMI system with MAHI Exo-II exoskeleton for stroke rehabilitation. A) In the schematic as presented in (Bhagat et al., 2020), the patient is trying to reach a target on the screen using the motor intent (monitored using EEG) and the attempt (monitored using EMG from the biceps and triceps) which in turn initiates the movement of the exoskeletonHaving a successful trial implied the involvement of fronto-parietal and cingulo-opercular networks and at the same time, the EMG signals went beyond a threshold confirming the attempt. B) The protocol timeline used in the study. C) Regions in the Fronto-Parietal Network (FPN) based on the DK atlas. D) Regions in the Cingulo-opercular Network (CON) based on the DK atlas. E) To move the exoskeleton, the patients have to both intent and attempt, which were monitored using the EEG and EMG signals respectively.

To monitor the efficiency of the protocol in improving motor ability, all the patients were assessed using the Fugl-Meyer Assessment for Upper Extremity (FMA-UE) and the Action Research Arm test (ARAT) at various timepoints during the study: baseline, within one week post-therapy completion, at 2 weeks follow-up, and 2 months follow-up (**Fig. 1B**). In our current analysis-focused study, we considered the scores during baseline and post 12-sessions of intervention. The FMA-UE test has 8 scoring sections assessing the movements involving flexor synergy, extensor synergy, combined synergies, out-of synergies, hand, wrist, coordination, and reflexes (Sullivan et al., 2011). The ARAT scores assess fine movements like grasp, grip, pinch, and gross movements (Yozbatiran et al., 2007). Our present work set out to analyze EEG, EMG, and MRI data further to understand the neural correlates of the associated stages of motor movement and its recovery.

### 2.2. EEG & EMG preprocessing

The original 64-channel dataset consisted of 56 EEG channels and 8 peripheral channels that were used to measure four differential EMG signals from the biceps and triceps muscles of both arms. The basic preprocessing of the EEG data consisted of zero-phase filtering (using a bandpass FIR (0.5 to 40 Hz) filter with an order of 1000), downsampling from 500 Hz to 250 Hz, and removing and interpolating the bad channels. On average, 2.4 ± 3.7 channels were removed per subject, with a median of 0.5 channels removed. Most of the data were of high quality hence, the majority of subjects had fewer than 3 channels removed.

This was followed by performing an Independent Component Analysis (ICA) using the Infomax algorithm (in EEGLAB (Delorme & Makeig, 2004)) with the default parameters to remove eye, muscle, and line noise artifacts. The data were then band-pass filtered into theta (3 to 7 Hz) and beta (13 to 30 Hz) bands, based on their relevance to our study. A custom MATLAB script (R2023b, The MathWorks, Inc.), incorporating EEGLAB functions, was used to preprocess all 12 sessions for each participant. To simplify the analysis, we grouped the 12 sessions’ data into 3 broader timelines (or SessionGroups) each comprising 4 individual sessions (i.e. new session 1 contained averaged data from sessions 1-4, new session 2 contained averaged data from sessions 5-8, and so on). The SessionGroups are further termed as just sessions in the rest of the manuscript, which are projected as the average of individual sessions within the group.

Raw EMG signals are bandpass filtered (30 – 200 Hz, 4^th^ order Butterworth) and then denoised using the Teager-Kaiser energy operator, followed by calculation of EMG envelope using a low-pass filter (0.5 Hz, 4th order Butterworth). The resulting data is standardized and compared against a threshold of 0.5 standard deviations to identify intervals of either flexor or extensor contraction. Contraction intervals larger than 1 second are retained for further analysis and their time of onset is utilized are marked as movement “attempt” events.

### 2.3. Segmenting EEG data based on trial outcomes

We segmented the EEG data for four trial outcome conditions: Success, Intention-only, Attempt-only, and Rest. As shown in **Fig 2A**, a successful trial condition occurs when an EEG-based intention is followed by an EMG-based attempt within 1 second, leading to the successful onset of the exoskeleton’s movement. If the interval between intention and attempt markers is beyond one second, then the trial is unsuccessful and does not trigger the exoskeleton’s movement as shown in **Fig. 2B**. **Figures 2C & 2D** present examples of trials when only one type of event occurs i.e. either intent only or attempt only conditions.

**Fig. 2.**
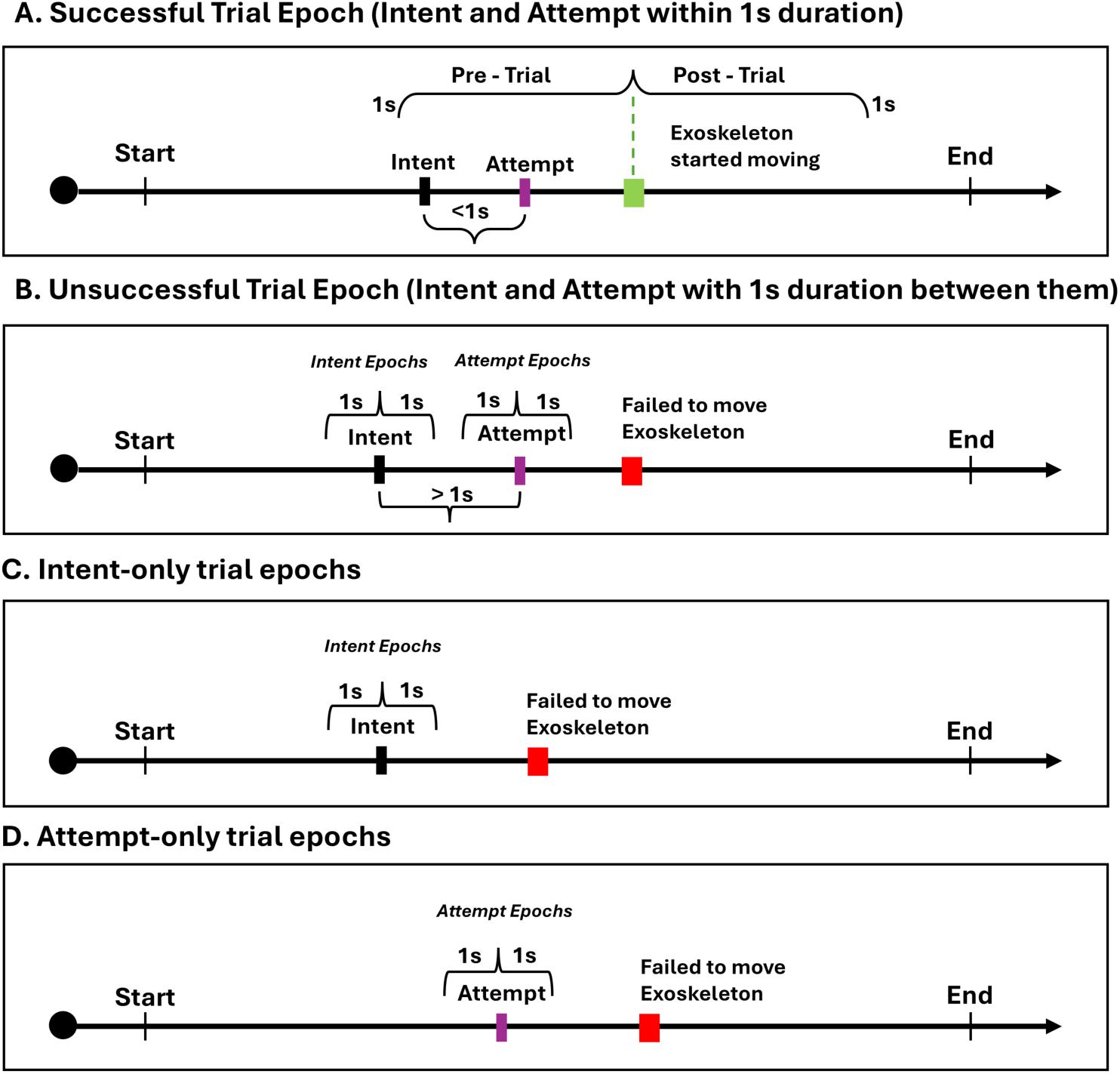
Illustration on the types of epochs. A) The successful trial epoch, in which intention and attempt signals were detected within 1s duration, and the exoskeleton successfully started moving. B) Unsuccessful trial epoch where intent and attempt were present but duration within was more than 1s. C) Another example of an unsuccessful trial epoch where only intent was present. D) Or only attempt was present

For extracting epochs for each condition, we used the different event markers that were time-locked to specific events above. Each epoch is 2 seconds long (1 second before and 1 second after the respective marker, called *pre* and *post-* in the rest of the manuscript). As both the intention-only and attempt-only trials constitute unsuccessful trials, we used the markers that reflect the presence of ‘attempt’ (from EMG) and ‘intention’ (from EEG) as the center point for the 2-second epochs. The Rest epochs were extracted from the resting period (40 seconds) at the beginning of each session. We extracted epochs separately for both the theta and beta band signals and therefore, we have 2 bands (theta and beta), 3 sessions, and 4 conditions constituting 24 trials in total for each patient.

### 2.4. MRI acquisition

The study used a 3T Ingenia (Philips) full-body MRI scanner to obtain each patient’s T1-weighted Magnetic Resonance Images (MRI). The scan protocol had the following acquisition parameters: number of acquisitions = 1; acquisition matrix = 252 × 227; TR = 8 ms; field of view = 250 × 200; duration = 5 min, 30 s; slice thickness = 2 mm; flip angle = 8 °; reconstructed in-plane resolution = 0.78 mm. For more information on this, please refer to (Bhagat et al., 2016).

### 2.5. Source estimation

We used the Recursive Spatial Bayesian Learning (RSBL) algorithm (Ojeda et al., 2018), which isolates independent components within the brain by imposing prior anatomical information and sparsity constraints, to localize the cortical sources according to the Desikan-Killiany (DK, 68-brain regions) atlas (Desikan et al., 2006). First, the MRI files were converted from Dicom to Nifti data format, imported into the Brainstorm (Tadel et al., 2011) and then the fiducial points were marked. Next, using CAT12 (Gaser et al., 2024), we segmented the MRI data into the scalp, skull, and brain. We generated 8003 vertices for the cortex surface. Then, we moved on to the next step of generating triangular meshes for each surface using the BEM method (Q. Liu et al., 2004). The number of vertices per layer was as follows: Scalp: 1082, Skull: 642, and Brain: 642, and a skull thickness of 4mm was assumed. We imported the EEG channel file and co-registered the electrode positions to the generated head model for each patient. Then, we computed the lead field matrix using Openmeeg (Gramfort et al., 2010). Finally, using a custom MATLAB script, we gathered the files generated from Brainstorm to create another head model (.mat) file that is compatible with the pop_rsbl function.

### 2.6. Analysis

#### 2.6.1. Network Analysis

We examined the activations and co-activations mediating the brain’s dynamical network states, specifically focusing on two networks: the frontoparietal network (FPN) and the cingulo-opercular network (CON). FPN includes the following regions of the DK atlas: caudalmiddlefrontal L, inferiorparietal L, parsopercularis R, precuneus L&R, rostralmiddlefrontal L, superiorparietal L&R, and supramarginal R where R and L stand for right and left respectively (**Fig. 1C**). The FPN focuses specifically on executive functions (Zanto & Gazzaley, 2013) supporting motor outputs and higher-order cognition (Olafson et al., 2022). As for CON, we included the following DK regions: superiorfrontal L&R, insula L, lateralorbitofrontal R, rostralmiddlefrontal L&R (**Fig. 1D**), which is implied in functions like task set maintenance or policy control, for perceptual readiness in maintaining alertness throughout a task (Hausman et al., 2021). Using RSBL, we computed the power time series for each network’s region of interest (ROI).

#### 2.6.2. Connectivity Analysis

We used the network activations through time for computing the directional connectivity quantified using the Transfer Entropy measure. We used the Java Information Dynamics Toolkit (JIDT) (Lizier, 2014) to compute Kraskov’s estimate of transfer entropy (Kraskov et al., 2004). It quantifies how much uncertainty has been reduced in predicting a series Y when using the past information of another series X. The amount of uncertainty reduced reflects the strength of the directional connectivity between the series. It is conceptually given by,

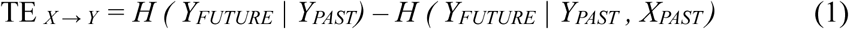

H is the Shannon Entropy. Higher values of TE imply more reduction in uncertainty for predicting Y with the help of X’s past (Vicente et al., 2011).

#### 2.6.3. Statistical Analysis

We were interested in observing how the effective connectivity between the FPN and CON changes over time in various spectral bands for those patients who improved functionally over time versus those patients, whose functional outcomes did not change from baseline. Specifically, we construct a repeated measures ANOVA (2 x 3 mixed design, **Table 1**) with two within-subject factors: Band (theta and beta) and Session (3 in number), and three between-subjects factors: Trial outcome (success, intention-only, attempt-only, and rest), subject group categorized based on gross motor improvement exhibited due to intervention (based on FMA), and that of fine motor improvement (based on ARAT). We asked if these factors could relate to the transfer entropy values of FPN and CON networks. We median split the FMA and ARAT score differences between the baseline and immediately post-treatment to classify patients into ‘improved’ and ‘not-improved’ subject groups. We are calling these groups: SubjectGroupFMA and SubjectGroupARAT (the value is 1 if shown significant improvement and 0 otherwise). For all epochs, we estimated the effective connectivity in both the pre and post-end of trial event (first half and second half) expecting to understand how these measures vary before and after a particular event marking the end of the trial (Success, Intent, Attempt, or Base). So, we had two models: one for the pre-event (FPN-CON Pre) and the other for the post-event (FPN-CON Post, **Table 1**).

**Table 1.**
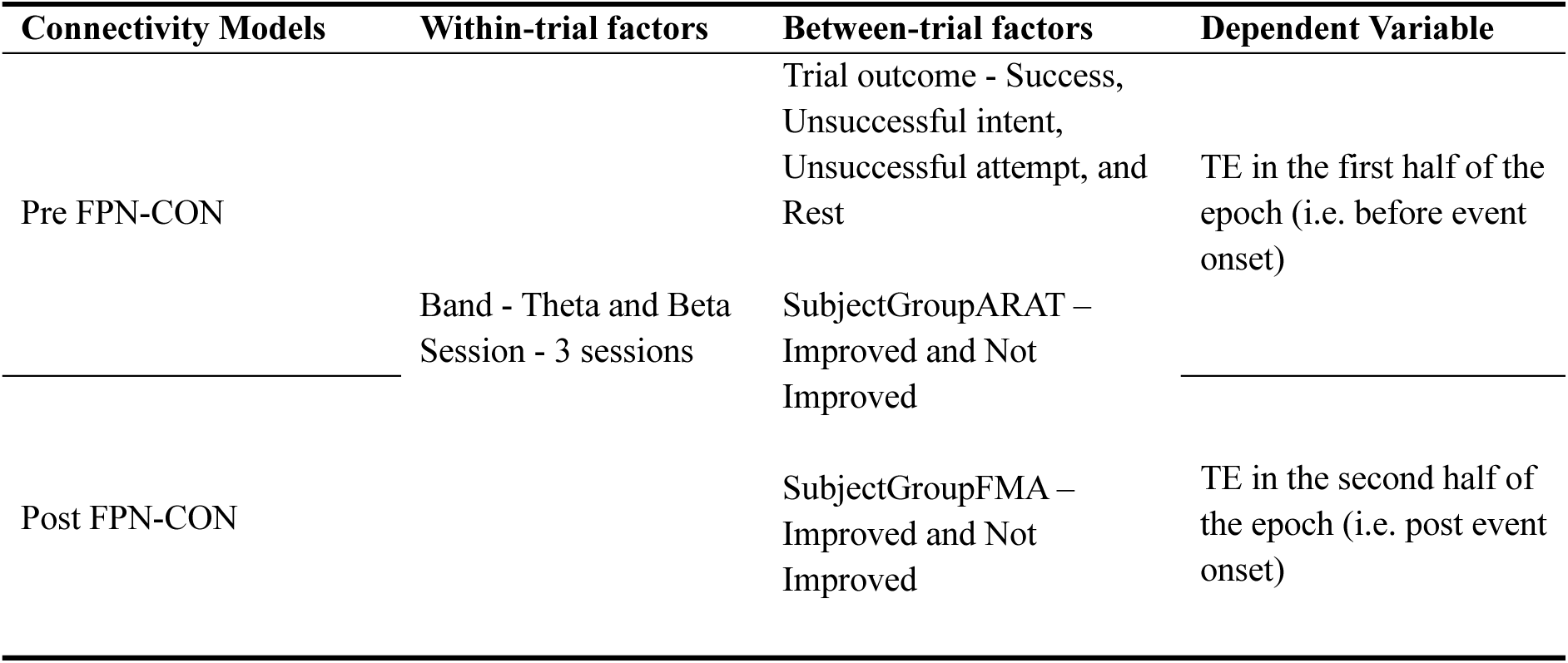
The table shows the variables used in the repeated measures ANOVA model. The SubjectGroupFMA and SubjectGroupARAT are categorized based on the median scores from each scale.

The statistics in the results are reported after correcting for sphericity which is the assumption that the relationship between any two levels of the repeated measures factor is homogenous across all pairs of levels. Mauchly’s test (Mauchly, 1940) using *mauchly* function (in MATLAB) indicated that the assumption of sphericity was violated for the models. Therefore, degrees of freedom were corrected and we report the Greenhouse-Geisser corrected p values.

#### 2.6.4. Regression Analysis

We also further abstracted the above model to a binary logistic regression model to specifically investigate the relationship between predictor variables (from the connectivity and the source power measures) and a binarized outcome variable (success or unsuccess where both unsuccessful intent and unsuccessful attempt trials constitute the ‘unsuccess’ trials). First, we assessed the importance of eleven predictors that consist of the connectivity between FPN and CON in pre and post-events, the pre-event and post-event FPN source power, pre-event and post-event CON source power, Age, Band (theta or beta), baseline ARAT and FMA scores, and finally the number of months since the stroke on predicting success vs unsuccess using a *stepglm* function. Then we cross-validated the optimum model using MATLAB’s *fitglm* function with a binomial distribution and a logit link function (**See Results 3.4**). The connectivity and power measures were baseline-corrected with the values from the Rest epochs for this analysis to predict the upcoming trial outcomes for any subject.

## 3. Results

Our results are organized to answer two main questions. We hypothesized that Fronto-Parietal and Cingulo-Opercular networks are critical for executive control and evaluation, and the effective connectivity between them and their source powers would explain patient-wise differences in presentation throughout the study. We first asked whether the involvement of FPN and CON networks would be different across the four different trial outcome conditions (successful trial, intent-only trials, attempt-only trials, baseline), and sought to identify significant substrates explaining the group differences (patients showing improvement versus otherwise). Next, we tested out the predictive utility of the identified substrates, we sought to know whether the networks’ power and connectivity measures are predictive of successful versus unsuccessful BMI decisions.

### 3.1. The pre-FPN-CON connectivity in the beta band can differentiate successful BMI trials, and a higher theta connection is related to gross motor improvement

To answer the first question, we set up a repeated measures ANOVA to understand whether the FPN and CON network connectivity measures are related to categories such as trial outcome, sessionGroup (blocks of sessions, see Methods), neural spectral band (theta or beta band), SubjectGroupARAT (ARAT based classification of patients improvement over time into improved (mean = 8.8, SD = 2.39) and not-improved groups (mean = 1.6, SD = 2.07)), and SubjectGroupFMA (improved: mean = 6.25, SD = 1.5; not-improved: mean = 1.67, SD = 2.34).

In the Pre FPN-CON model (i.e., connectivity measured using EEG signals from 1 second before an event occurred), there was a significant main effect of *trial outcome* (F(corrected degrees of freedom in the numerator = 1.82, corrected degrees of freedom in the denominator = 20.60) = 15.28, p < 0.0001, 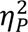 = 0.57), *Band* (F(0.61, 20.60) = 122.69, p < 0.0001, 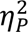 = “ ” 0.78) and a significant interaction between *Band* and *trial-outcome* (F(1.82, 20.60) = 4.00, p = 0.015, 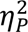 = 0.26) and between *SubjectGroupFMA* and *Band* (F(0.61, 20.60) = 5.72, p = 0.022, 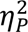 = 0.14). Multiple pairwise comparisons were conducted using the Tukey-Kramer test. The connectivity in the successful trials is higher compared to both attempt-only trials (mean difference = 0.071 ± 0.016, p < 0.001) and rest (mean difference = 0.096 ± 0.016, p < 0.0001) conditions. Similarly, intent-only trials showed significantly higher connectivity than attempt-only trials (mean difference = 0.049 ± 0.016, p = 0.02) and rest (mean difference = 0.074 ± 0.016, p < 0.001) (**Fig. 3A**). No significant differences were found between the connectivity in successful and intent-only trials (p = 0.528) or between attempt-only and rest (p = 0.413). Moreover, the connectivity in the theta band was significantly stronger than the beta (mean difference = 0.118 ± 0.011, p < 0.0001, **Fig 3B**), however, the connectivity in the successful trials was significantly differentiable from all other trial outcomes in the beta frequency band (Success vs Intent-only: mean difference = 0.02 ± 0.009, p = 0.034; Success vs Attempt-only: mean difference = 0.04 ± 0.009, p < 0.001; Success vs Rest: mean difference = 0.06 ± 0.009, p < 0.0001). The interaction between the band and trial outcome is shown in **Fig. 3C**.

**Fig. 3.**
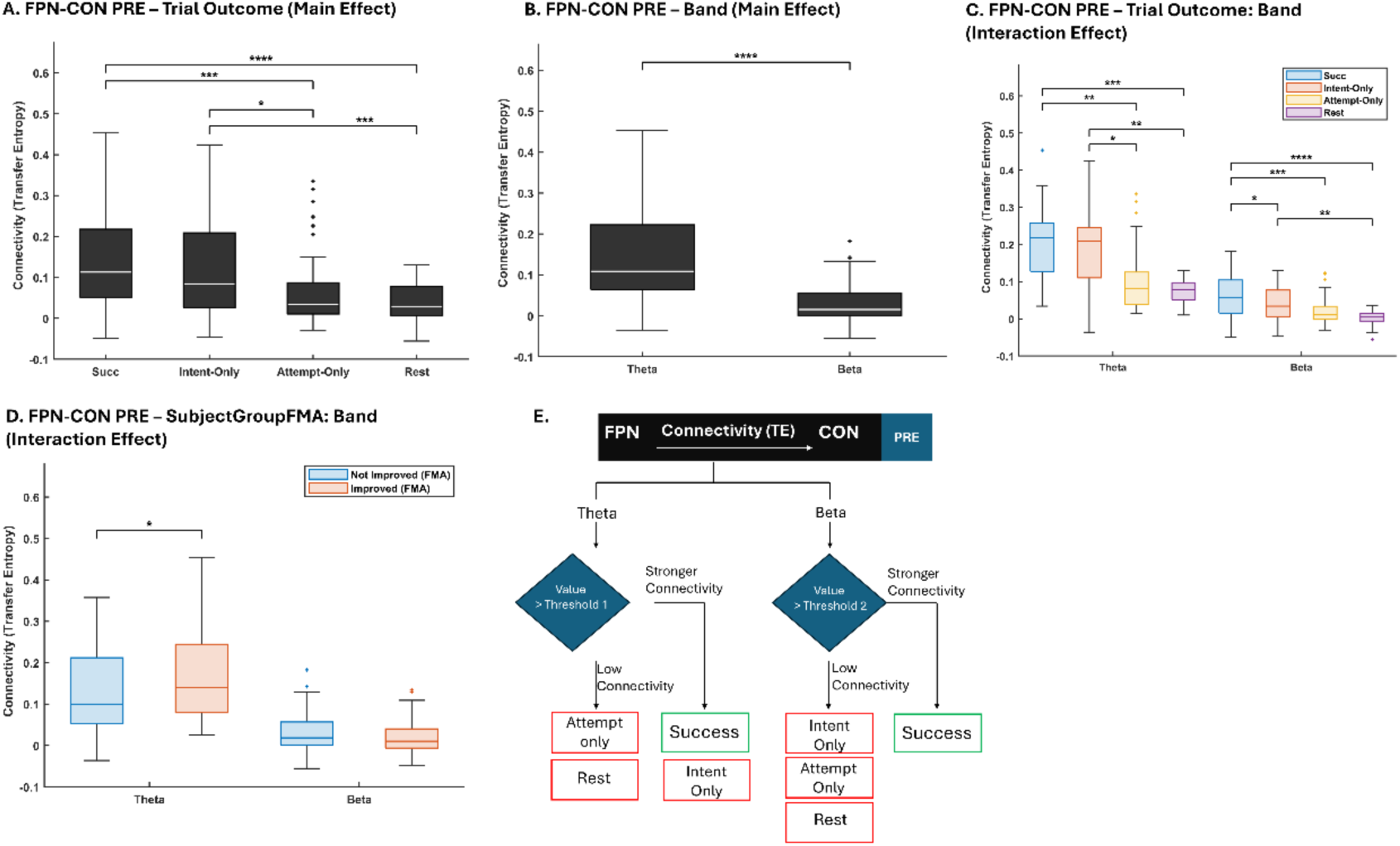
The post hoc results from the FPN-CON Pre model. A) Differences in TE among the 4 trial outcome conditions. B) The overall connectivity is stronger in the Theta band than in the Beta. C) The boxplot shows how the TE for each trial outcome was different in the two bands. D) Improved group based on the FMA scores showed stronger theta connectivity. E) A summary schematic showing the interaction effect. Strong beta connectivity between FPN-CON before the trial leads to a successful trial completion (moving the exoskeleton). Since there was no significant difference between successful and unsuccessful intent in theta, the same conclusion can’t be reached. *p<0.05, **p<0.01, ***p<0.001, ****p<0.0001

Interestingly, we have observed a significant pre-event connectivity difference between improved and not-improved groups based on the FMA score in the theta band. Improved groups showed stronger theta connectivity compared to the not-improved group (mean difference = 0.04 ± 0.02, p = 0.045) (**Fig. 3D**). The groups categorized by the ARAT scores did not show any significant differences in connectivity (p = 0.301).

### 3.2. The post-FPN-CON connectivity in the beta band can also differentiate successful BMI trials and a lower theta connection is related to fine motor improvement

Similar to Pre FPN-CON Pre, the Post FPN-CON model (i.e., connectivity measured using EEG signals from 1 second after an event occurred) also showed a main effect of *trial outcome* condition (F(1.92, 21.75) = 9.12, p < 0.001, 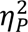 = 0.44), *Band* (F(0.64, 21.75) = 70.44, p < 0.0001, 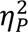 = 0.67) and a significant interaction between *Band* and *Condition* (F(1.92, 21.75) = 3.53, p = 0.025, 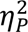 = 0.24). Tukey-Kramer test on condition main effect showed that all three conditions were significantly different from the rest (Success vs Rest: mean difference = 0.089 ± 0.018, p < 0.001; Intent-only vs Rest: mean difference = 0.074± 0.018, p = 0.002; Attempt-only vs Rest: mean difference = 0.05 ± 0.018, p = 0.049) **(Fig. 4A)**. Again, the connectivity in theta band was stronger compared to the beta (mean difference = 0.09 ± 0.011, p < 0.0001), however, the connectivity in successful trials was significantly differentiable in the beta frequency band (Success vs Intent-only: mean difference = 0.038 ± 0.012, p = 0.012 Success vs Attempt-only: mean difference = 0.034 ± 0.012, p < 0.031; Success vs Rest: mean difference = 0.062 ± 0.012, p < 0.0001, **Fig. 4C**).

**Fig. 4.**
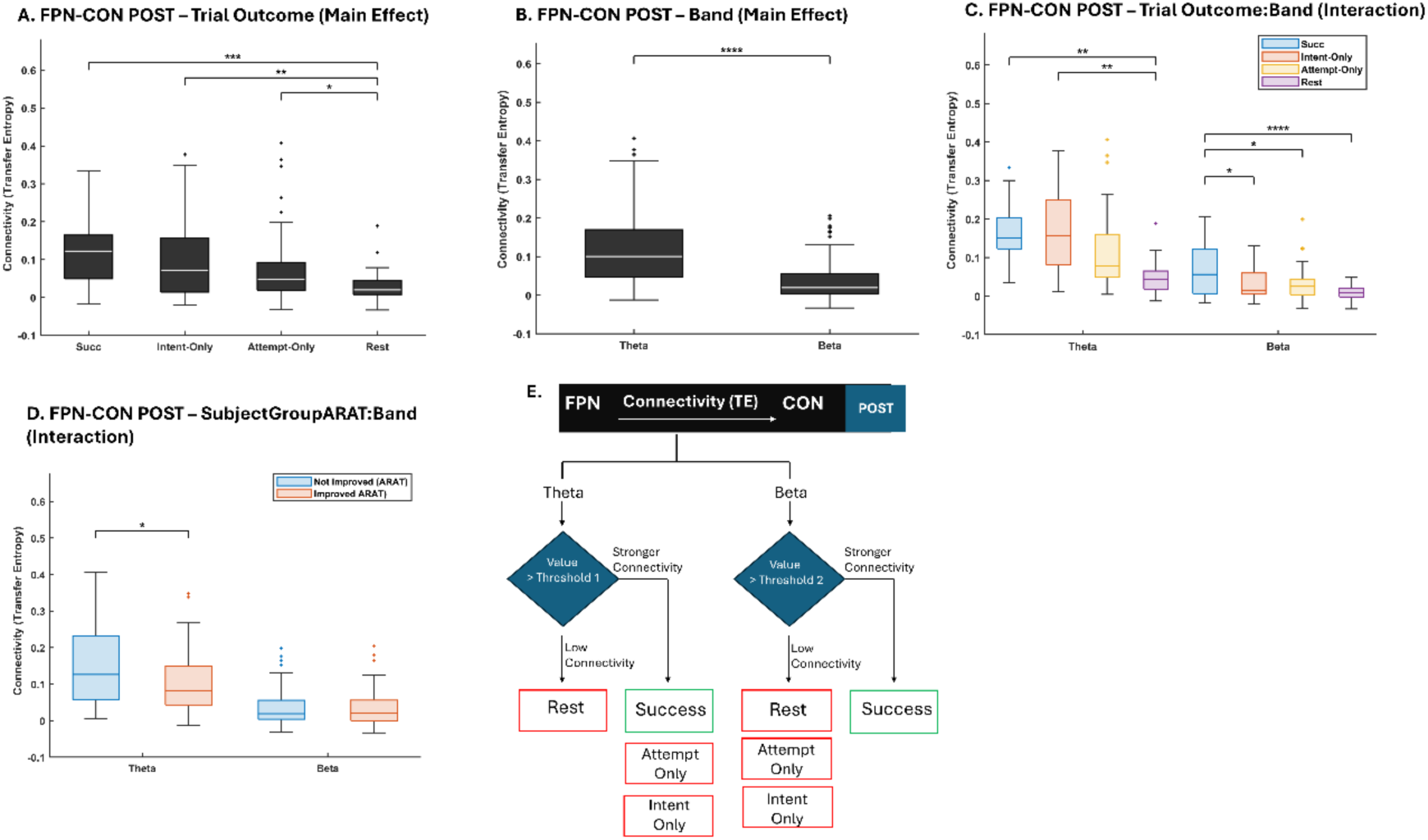
The post hoc results from the FPN-CON Post model. A) Trial outcome main effect. B) Band main effect. C) Interaction between trial outcome and band. D) Interaction between the band and SubjectGroup ARAT (improved vs not improved groups). E) Schematic showing the summary of Band and Trial outcome conditions in the FPN-CON Post model. Strong connectivity in beta for the FPN-CON after an trial can lead to a successful trial. In theta, there were no significant differences between Success, Unsuccessful Intent, and Unsuccessful Attempt. *p<0.05, **p<0.01, ***p<0.001, ****p<0.0001

In addition, we observed an interaction between *SubjectGroupARAT* (improved group vs not improved based on ARAT) and *Band* (F(0.64, 21.75) = 4.14, p = 0.049, 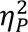 = 0.11), and it was the theta band found to be significantly lower in those who improved over time (mean difference = 0.048 ± 0.022, p = 0.042, **Fig. 4D**). However, the SubjectGroupFMA (improved group vs not improved based on ARAT) has not shown any interaction or main effects in the model (p = 0.733).

### 3.3. Spectral Power of the FPN and CON networks align with the connectivity results

We also computed the source power values of the FPN and the CON networks in the pre and post-event for the epochs corresponding to each trial outcome. Repeated measures ANOVA model was fitted with source power as the dependent variable and with Band and Sessions as within-subject factors and Condition, SubjectGroupFMA, and SubjectGroupARAT as between-subject factors (Please see the Supplementary for detailed analysis). **Table 2** shows the significant variables for four models investigating dependent variables as powers of different sources (Pre and post-FPN and CON networks).

**Table 2.**
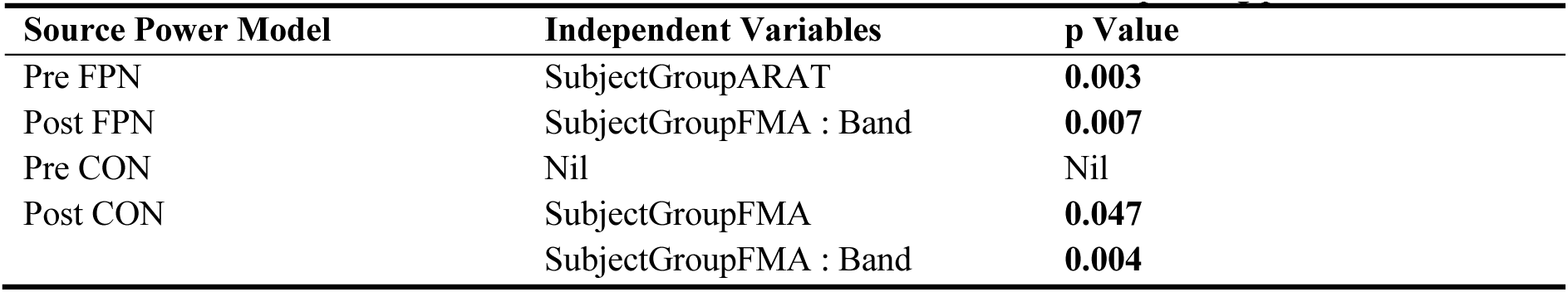
The table shows the main and interaction effects from each model and the corresponding p values

The FPN power in the pre-trial time period significantly differed between the groups based on ARAT (**Fig. 5A**, ARAT: F(0.48, 11.95) = 10.72, p < 0.003, 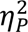 = 0.3) where the improved group showed lesser (more desynchronized) power than the not-improved group (mean difference = 0.006 ± 0.002, p = 0.003), whereas the Pre-CON Power model did not show any significant relation to improvement. Interestingly, the Post-CON model showed a main effect of SubjectGroupFMA (F(0.41, 10.22) = 4.37, p = 0.047, 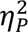 = 0.15) and an interaction between SubjectGroupFMA and Band (F(0.41, 10.22) = 9.95, p = 0.004, 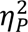 = 0.28). In theta band, the patient groups based on the FMA scores significantly differed in which the improved group showed higher CON power (synchronization) in the post-trial event time period compared to the not-improved group (mean difference = 0.03 ± 0.012, p = 0.016, **Fig. 5B**). Moreover in the post-trial FPN Power model, we observed an overall significant difference between the two bands (mean difference = 0.012 ± 0.003, p < 0.001) where the theta band power was larger than the beta power in the improved group based on FMA scores.

**Fig. 5.**
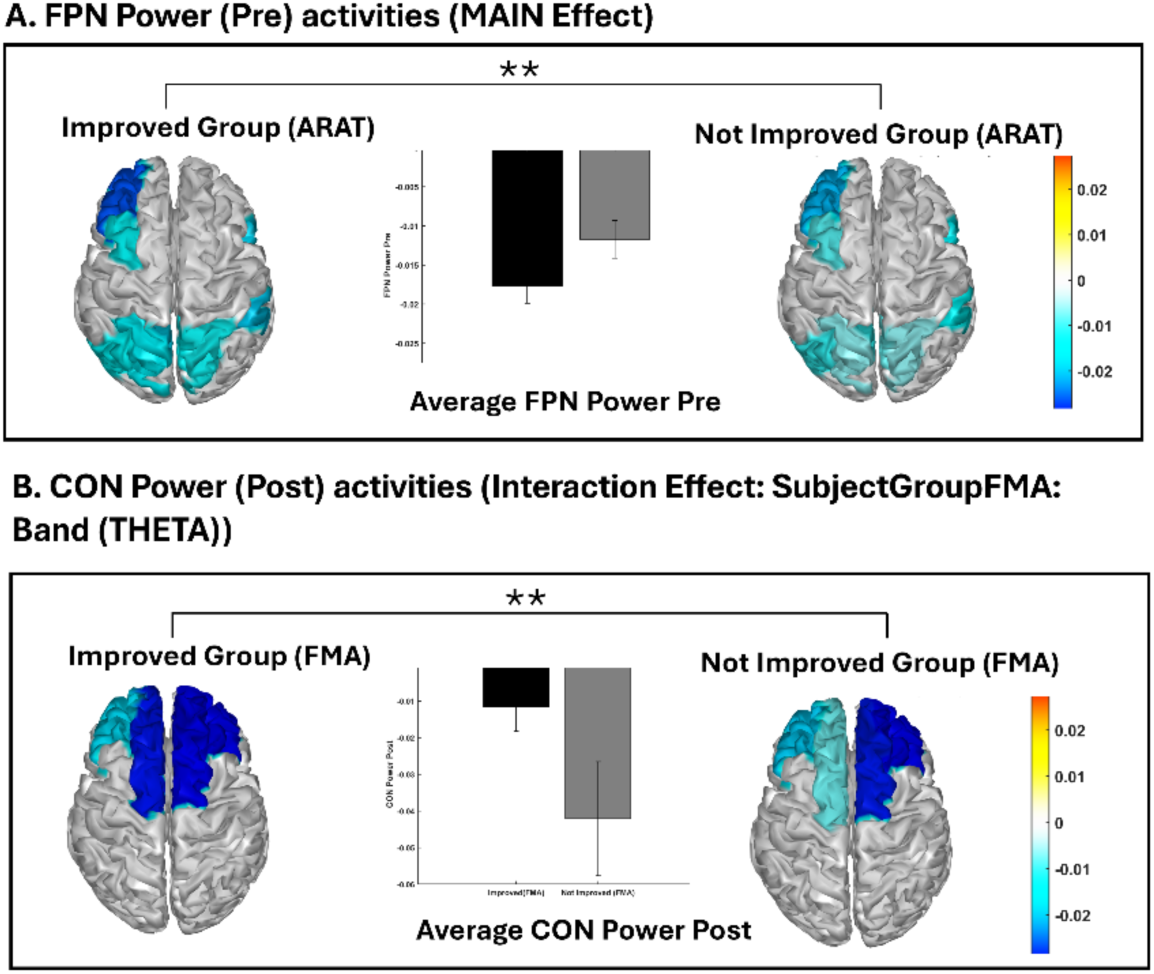
A) The activities (broadband) over the cortical surface for FPN are shown for the groups categorized based on ARAT scores suggesting higher desynchronization in the improved group. B) The activities in the theta band over the cortical surface for CON are shown for the groups categorized based on FMA scores suggesting higher desynchronization in the not-improved group. *p<0.05, **p<0.01, ***p<0.001, ****p<0.0001

### 3.4. Connectivity and source power measures are significantly predictive of trial success

To understand the predictive utility of the successful trial network activations, we set up a stepwise binary logistic regression model (*stepwiseglm* function in MATLAB) with eleven predictor variables and selected the significant predictors based on deviance as the criterion (small deviance implies that the prediction is closer to the observed outcomes). The eleven predictors consist of the connectivity between FPN and CON in pre and post-trial events, the pre and post-FPN source power, the pre and post-CON source power, Age, Band (theta or beta), baseline ARAT and FMA scores, and finally the number of months since stroke (See demographics of patients in the supplementary). The connectivity and power measures were baseline corrected using those assessed in the resting period, and the outcome variable was binarized categorizing the 3 trial outcomes as either 1 (success) or 0 (Intent-only and Attempt-only). All predictors were z-score normalized. The optimal model after the stepwise procedure had the following predictors: connectivity between FPN and CON in the pre and post-trial and their interaction, power measures of post-FPN and pre-CON, and their interaction (**Table 3**). The model (10 patients x 2 bands x 3 trial outcomes x 3 sessions, therefore 173 degrees of freedom for error) showed strong statistical significance compared to the constant model (χ²(7) = 43.7, p < 0.0001.

**Table 3.** The estimated coefficients, standard errors, test statistics, and p-values for the logistic regression model

| Predictor | Estimate | SE | tStat | pValue |
| --- | --- | --- | --- | --- |
| Pre connectivity | 1.007 | 0.257 | 3.922 | < 0.0001 |
| Post connectivity | 0.394 | 0.229 | 1.717 | Ns |
| Post FPN Power | 0.237 | 0.240 | 0.985 | Ns |
| Pre CON Power | -0.824 | 0.385 | -2.138 | 0.032 |
| Pre:Post connectivity | -0.866 | 0.240 | -3.612 | < 0.001 |
| PostFpn:PreCon Power | -0.535 | 0.254 | -2.104 | 0.035 |

Higher FPN-CON Pre connectivity showed a significant relation to the success of a trial (β = 1.007 ± 0.257, p < 0.0001). However, the FPN-CON Post connectivity was not significant (p = 0.086), however, a significant negative interaction effect was observed between both predictors (β = -0.866 ± 0.24, p < 0.001). In the power measures, the pre-CON was significant (β = -0.824 ± 0.385, p = 0.032), however, the post-FPN Power was not (p = 0.324). And, there was a significant negative interaction between the post-FPN power and the pre-CON power (β = -0.535 ± 0.254, p = 0.035, **Table 3**). The model suggested that the pre-connectivity features were relatively strong predictors of trial success.

To better understand the negative interactions, we computed the likelihood of success at all possible pairs of the variables that showed the interactions and plotted a surf map (**Fig. 6C & D**). While changing the interaction terms, all other variables were kept constant at their mean values. In the interaction between pre and post-connectivity measures, the chance of success tends to be higher if one of the measures is higher and the other is lower. In other words, for a trial to be successful, strong pre-connectivity was necessary if and only if post-connectivity was lower. Out of the power measures that were significant, we observed similar negative interaction like that of the connectivity, in which either a strong post-FPN or pre-CON of a trial was required for a successful trial, with the other one being lower.

**Fig. 6.**
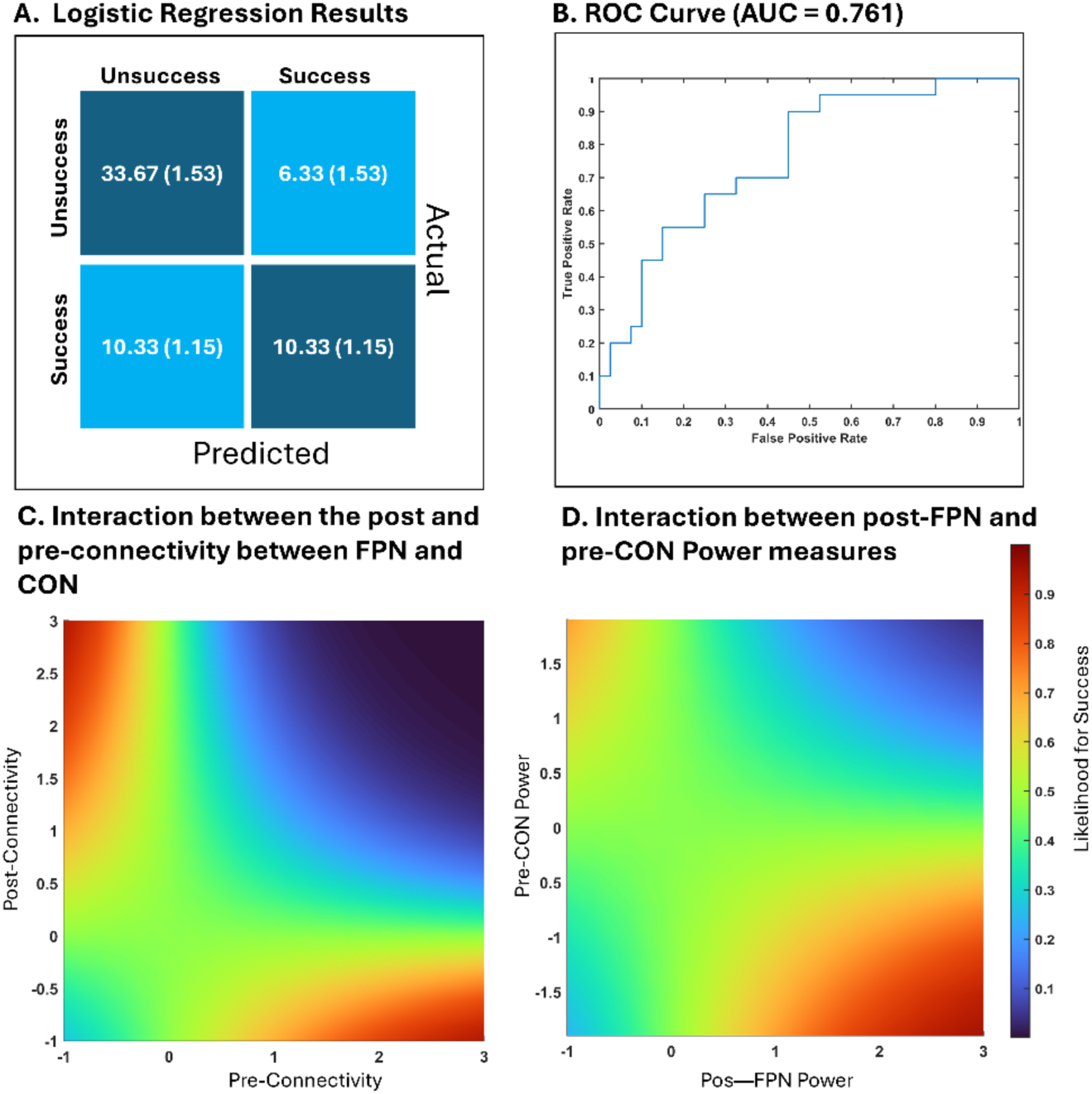
A) The confusion matrix showing the average from the 3-fold cross-validation for the binary logistic regression model, also suggests a larger specificity of prediction to about 84.2%. B) The ROC curve after cross-validation. C) Interaction between FPN-CON connectivity in the pre and post-trial event, suggesting a negative interaction can increase the chance of a successful trial. D) Similar negative interaction between post-FPN power and pre-CON suggestive of trial success. The color bar represents the likelihood of success, and the axes represent the zscored normalized units.

The model’s predictive performance was further assessed using 3-fold cross-validation with stratified training and testing sets **(Fig. 6A).** The average accuracy of the model was 0.722 (± 0.010). The average sensitivity and specificity were 0.483 (±0.058) and 0.842 (±0.038) respectively. The model achieved an AUC of 0.761 (**Fig. 6B**) and the average F1 score was 0.536 (±0.026).

## 4. Discussion

Recent studies suggest that the brain patterns during motor preparation can relate to the underlying motor intent for controlling limb movements. Moreover, a closed-loop control system that feeds in these neural patterns for controlling the limb can systematically assist in facilitating the circuit plasticity activations required for motor recovery. Similarly, understanding an individual’s error-related neural potential can assess the extent of learning happening in the patient, and inform about subjective attention, motivation, and active control exerted by the patient on any performance (Mitra et al., 2023; Soriano-Segura, Ferrero, Ortiz, et al., 2023). Not surprisingly, there has been recent evidence showcasing the importance of cognitive networks in motor recovery (Lee & Kim, 2023; Mattos et al., 2023). Recent reviews also point out the importance of looking at the motor strip for reward information as well (Derosiere et al., 2025).

In the current study, we particularly focussed on circuit-level phenomena mediating BMI control. It is the neural circuit involving many nuclei and their interactions that have been suggested to integrate information and holistically encode signals required for timed motor actions. Although many studies mentioned earlier focus on specific neural regions for decoding a cognitive or motor function, such as the motor cortex or the anterior cingulate cortex, those exploring the circuit-level activations for informing the upper limb rehabilitation performance aren’t well understood. The networks encompassing the motor cortex strip is the fronto-parietal network that encodes the motor preparation and executive information, while that encompassing the anterior cingulate cortices is the cingulo-opercular network that is suggestive of encoding the utility of the action. We hypothesize that the power and the connectivity between executive fronto-parietal networks (FPN) and reward process evaluating cingulo-opercular networks (CON) are informative of the success of the trial. The advantage of developing a BMI decoder based on the circuit activation rather than a focused region-based signal is its ability to represent the individual differences in signaling the phenomena more holistically. The network activations offer the ability to understand the motor-related planning and execution, sensory-motor integration, learning, and adaptation, of relevance to stroke rehabilitation (Arber, 2012; Cisek & Kalaska, 2010; Hanakawa et al., 2003a).

Many studies have correlated the cortical functional reorganization change in the resting state before and after a rehabilitation protocol to the recovery rate in stroke patients (Hordacre et al., 2020; Volz et al., 2016), and propose that post-stroke recovery depends on the brain’s capacity to restore interhemispheric connectivity. Many also focus on computing functional connectivity within/between many networks in stroke patients, especially the default mode network (DMN): DMN increases its activity at rest and the activity goes down when the brain is engaged in a goal-directed task (Menon, 2023), A review by (Ismail et al., 2024) suggest that functional connectivity within and between the default mode network is associated with cognitive recovery in post-stroke patients. While traditional assessment approaches give a measure of behavioral restoration (Rathee et al., 2019), we propose the functional connectivity-based neuro-physiological to precisely understand reward value encoding versus execution during the BMI task. We further present the possibility of using these connectivity measures to predict the trial outcome for any subject passively.

Our results provide several insights: The beta connectivity between FPN and CON was able to distinguish successful trials from the rest and the unsuccessful, in both pre and post-trial event conditions. The prefrontal beta power’s role in executive control of action has been suggested earlier (Schmidt et al., 2019), and our study proposes a possible circuit connectivity mechanism underlying beta oscillation for the successful execution of an action. Overall broader connectivity was the strongest for successful BMI trials and they primarily contributed to predicting the success with about 84.2% specificity. Motor imagery and execution demand differential connectivity patterns among regions involving the prefrontal cortex (Sharma et al., 2009). In our study, we used this measure as a marker to know and predict the cognitive state of the patient.

Second, increased theta connectivity in pre-trial can be informative of the extent of FMA-based score improvement to be expected in patients (higher theta connectivity is associated with improvement), while during the post-trial period can be informative of ARAT improvement (lower theta connectivity is associative with improvement). In terms of network power, more synchronized post-CON in theta band was reflective of success and FMA improvement. And, highly desynchronized pre-FPN was indicative of ARAT improvement. Moreover, we estimated the marginal effects of the logistic regression model to understand the negative interactions between connectivity and power measures. The marginal effect of pre-connectivity on the success of the trial was related to the state of post-connectivity to reflect success. That is, to complete the trial successfully, either pre or post-connectivity has to be higher, and not both. The same observation was found between power measures (pre-CON and post-FPN). This shows the importance of monitoring both the pre and post-trial connectivity and power measures to predict the BMI outcome while they undergo rehabilitation.

## Outlook and Future works

The quality of cognitive control is critical to performing our tasks independently and efficiently and maintaining interpersonal relationships. Approximately 2/3^rd^ of stroke patients experience cognitive decline and impairment, out of which over 50% remain in the same state of cognitive impairment after a year post-injury (El Husseini et al., 2023). Cognitive impairment or dementia after stroke is predominantly defined by dementia that occurs within three months after stroke onset. Many stroke survivors develop delayed dementia beyond three months or only after recurrent stroke(s). The recognition of cognitive impairment in the acute phase after stroke may offer vital information to the clinician for early cognitive rehabilitation (Kalaria et al., 2016). A review of the literature shows that the prevalence of post-stroke cognitive impairment (PSCI) varies from 6 percent (De Ronchi et al., 2007) with total subjects of 7930, 21 percent (Khedr et al., 2009), 37.1 percent (Chongqing Stroke Study, n= 434 (Zhou et al., 2005)), to 80 percent (Sun et al., 2014). A population survey conducted in Kolkata found that 13.9 percent of stroke survivors developed post-stroke dementia and 6.1 percent had mild cognitive impairment (Das et al., 2013). For patients with PSCI, attention, learning, memory, language, visuospatial/constructional functions, speed, frontal executive function, nominal skill, perceptual skill, visual memory, and verbal memory are among the cognitive functions that are commonly affected. Therefore, we need to come up with interventions that are accessible to cognitively impaired patients as well.

One possibility is to develop a closed-loop BMI system that passively assesses the underpinning neural patterns and precisely adjusts the cognitive difficulty of the rehabilitation paradigm for individual needs. The advantage of developing such a BMI decoder based on the circuit activation rather than a focused region-based signal is its ability to represent the individual differences in signaling the phenomena more holistically. Further, the network activations also offer the ability to understand motor-related planning and execution, sensory-motor integration, learning, and adaptation, which is highly relevant for stroke rehabilitation (Arber, 2012; Cisek & Kalaska, 2010; Hanakawa et al., 2003b)

Understanding the neural structures mediating the exact hierarchical stages underlying motor action is still an active area of research. Traditional BMI setups mostly relied on channel-space measures like the motor-evoked potential for monitoring intention and less explored the sensory-cognitive-motor network dynamics underlying the act. There are various reasons for this simplified approach, such as reducing the computational complexity of those processes in real-time. Our study took a further step by showcasing the richness of understanding the source space can provide about the higher-level stages like executive control, and motivation behind a movement and we reasoned that these measures can be used in traditional BMIs to enhance its overall efficacy. Yet, we are barely capturing the whole picture here. Considering a single successful trial in our paradigm, it may not be a simple intent-attempt pair that defines the whole trial. Other cognitive processes involving partial error corrections and stopping may have to be deployed to indicate the overall success of the trial. We propose the future BMI setups should target understanding these aspects to make it more friendly to the real-time patient’s needs.

Altogether, our study presents initial evidence for the circuit-level activations holding pragmatic information for modulating the BMIs, that can care for the personalized training with increased engagement of cognitively impaired stroke patients undergoing upper limb rehabilitation. One limitation is the computational complexity required for estimating the proposed neural measures in an online fashion for BMI control. In the future, we aim to address the real-time programming challenges and utilize the identified network markers of motor execution and its associated cognition to optimally adjust the cognitive load of any trial and facilitate learning of the trial outcome for accelerated recovery.

## CRediT authorship contribution statement

The study was conceived by NAB and PPB. NAB preprocessed the EEG and EMG data for source localization. GKRP and PPB performed the source localization and connectivity analysis. GKRP, PPB, and NAB contributed to the writing and reviewing of the manuscript.

## Ethics statement

The manuscript involves data analysis collected from an earlier conducted study. The original study procedures were approved by the Institutional Review Boards of University of Houston, Rice University, University of Texas Health Science Center at Houston, and the Houston Methodist Hospital at Houston, Texas. All participants provided informed consent in accordance with the Declaration of Helsinki.

## Acknowledgments

PPB thanks Dr. Nandini Priyanka for insightful discussions. All authors thank Drs. Subasree Ramakrishnan. Shantala Hegde and Navin B P, National Institute of Mental Health and Neuro Sciences, Bengaluru, India for the helpful discussions. We also thank the National Institutes of Health’s National Robotics Initiative (Grant #<u>R01NS081854</u>) and the TIRR Foundation (Mission Connect) for funding the data collection, which made this study possible.

## Data availability

Data from the current publication is available upon reasonable request to the corresponding authors - PPB, and NAB.

## Supplementary Material

### A. Patient Demographics

**Table 1.** Patient Demographics when presented during the start of the rehabilitation program

| Patient ID | Gender | In Age-range | Stroke type & location | Months Since Stroke | Paretic Arm | FMA - UE | ARAT |
| --- | --- | --- | --- | --- | --- | --- | --- |
| P1 | M | 70's | Hem., C | 72 | Right | 51 | 43 |
| P2 | F | 40's | Isch., C | 106 | Left | 26 | 4 |
| P3 | F | 50's | Isch., SC | 72 | Left | 48 | 45 |
| P4 | F | 50's | Isch., C | 14 | Left | 21 | 4 |
| P5 | M | 50's | Isch., SC | 10 | Right | 43 | 39 |
| P6 | M | 60's | Hem., C & SC | 9 | Right | 45 | 30 |
| P7 | M | 40's | Hem., C & SC | 21 | Right | 38 | 25 |
| P8 | M | 50's | Hem., C | 38 | Left | 49 | 42 |
| P9 | M | 60's | Isch., SC | 15 | Left | 20 | 9 |
| P10 | M | 40's | Hem., SC | 17 | Right | 37 | 12 |

Stroke type = Hem. = Hemorrhagic stroke, Isch. = Ischemic stroke
Lesion Location – C = Cortical, SC = Subcortical
FMA-UE = Fugl-Meyer Upper Extremity (0-66), ARAT = Action Research Arm Test (0-57)

### B. Channel level analysis

We were also interested in seeing if the channel powers corresponding to the two networks show any distinguishable activations for the conditions where the baseline activity is expected to be the minimum. For the FPN channels, we focused on F3, F4, Fz, FC1, FC2, FC3, FC4, P3, P4, Pz, CP1, CP2, CP3, and CP4 while Fz, FCz, F3, F4, FC1, FC2, Cz, C3, C4, and CPz formed the CON channels. The power spectral density (PSD) was calculated using Welch’s method. A segment length of 250 samples was used and the overlap was set to 75% of the window length. The PSD for each epoch for each condition in two bands was estimated for all the sessions and the subjects. The averaged values were plotted using a bar graph (**Fig. S1**).

**Fig. S1.**
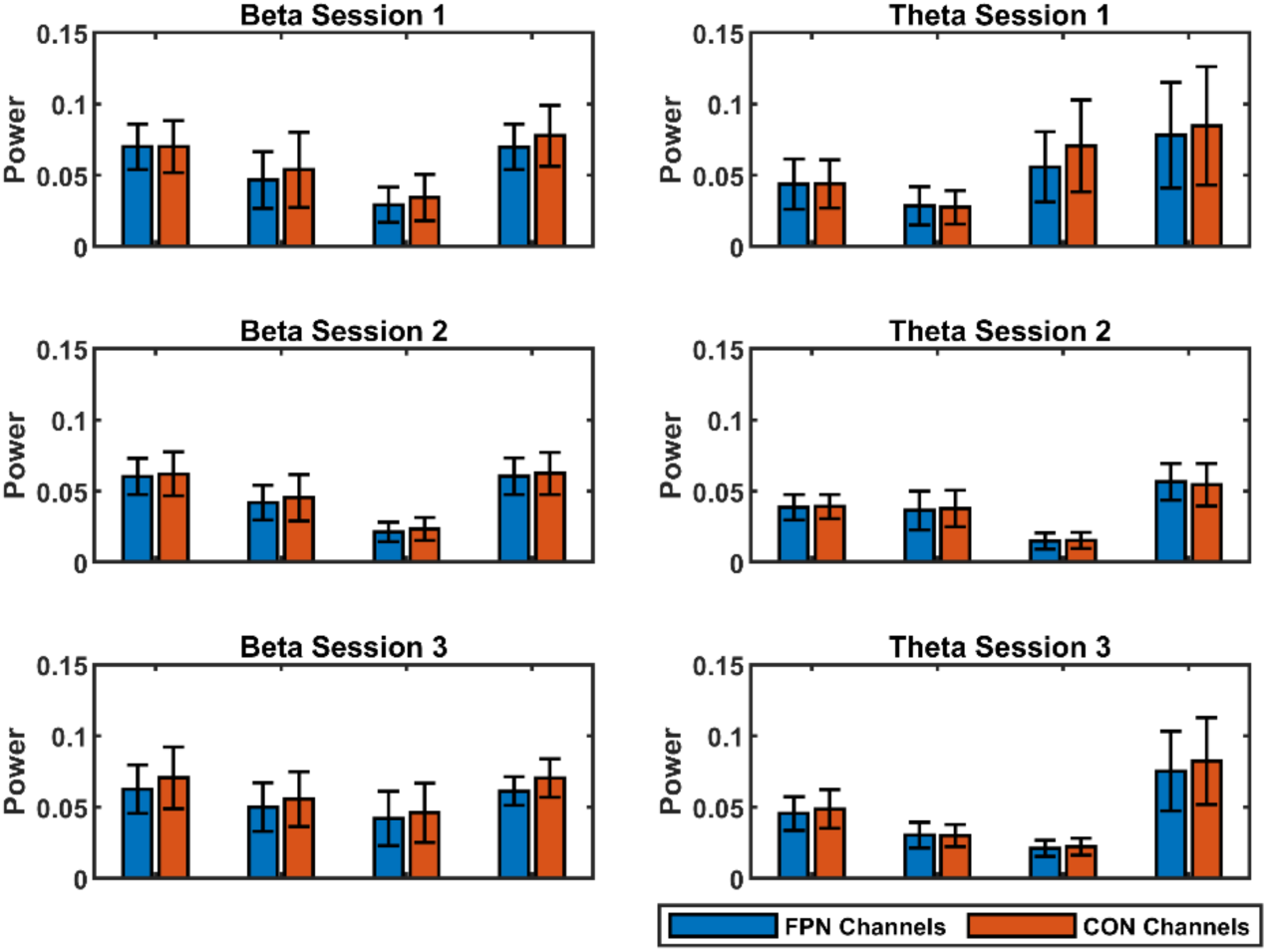
Channel level analysis. Average channel level powers in both FP and CO networks. Left: in theta band. Right: in beta band. Errorbars represent SEM.

### C. Source Power Analysis

Since the FPN-CON connectivity was a significant measure in predicting the success vs unsuccess trials, we were also interested in seeing how the source power changes among the conditions. We extracted the power values of the regions that form the FP and CO networks. Based on the DK atlas (Desikan et al., 2006)(Desikan et al., 2006), FPN is formed out of caudalmiddlefrontal L, inferiorparietal L, parsopercularis R, precuneus L, precuneus R, rostralmiddlefrontal L, superiorparietal L, superiorparietal R and supramarginal R. The CON has the regions superiorfrontal L, superiorfrontal R, insula L, lateralorbitofrontal R, rostralmiddlefrontal L, and rostralmiddlefrontal R. We used a repeated measures ANOVA to understand whether the FPN and CON power measures are related to categories such as trial outcome, sessionGroup, neural spectral band, and finally the patient improvement group. We conducted 4 separate analyses with different dependent variables: FPN power before the event marking end of the trial (Pre-FPN power), FPN power post the trial end (Post-FPN power), similarly for pre and post CON power (**Table 2**).

**Table 2.**
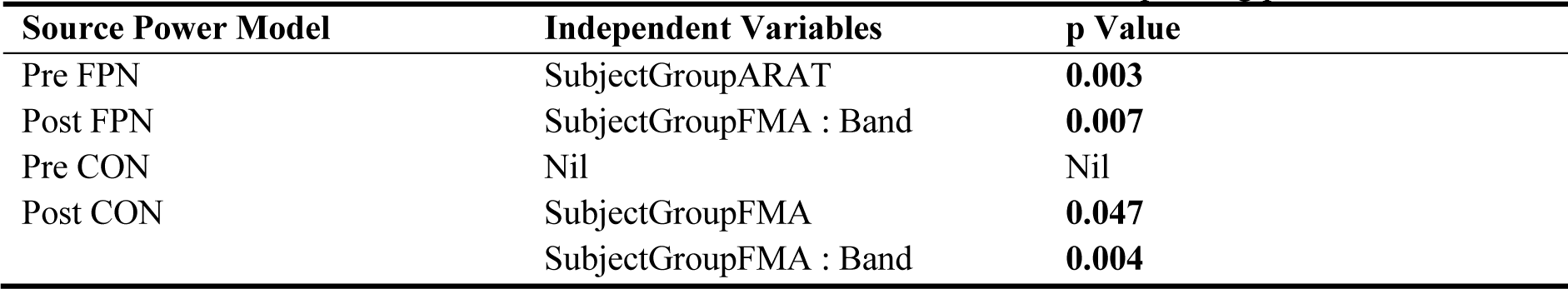
The table shows the main and interaction effects from each model and the corresponding p values

